# Surveillance of *Neisseria meningitidis* Carriage Four Years After menACWY Vaccine Implementation in the Netherlands Reveals Decline in Vaccine-type and Rise in Genogroup E Circulation

**DOI:** 10.1101/2023.02.24.23286220

**Authors:** Willem R. Miellet, Gerlinde Pluister, Meike Sikking, Marcia Tappel, Jurgen Karczewski, Linda J. Visser, Thijs Bosch, Krzysztof Trzciński, Rob Mariman

**Author notes:** **Correspondence:** Rob Mariman. These authors contributed equally to this work and share last authorship (Krzysztof Trzciński, Rob Mariman).

## Abstract

Carriage of *Neisseria meningitidis* is an accepted endpoint in monitoring meningococcal vaccine effects. We applied molecular methods to assess the impact of menACWY vaccine implementation on meningococcal carriage and genogroup-specific prevalence in young adults in Fall of 2022, four years after the introduction of the tetravalent vaccine in the Netherlands. The overall carriage rate of genogroupable meningococci was not significantly different compared to the pre-menACWY cohort investigated in 2018 (20.8% or 125 of 601 versus 17.4.% or 52 of 299 individuals, *p*=0.25). Of n=125 carriers of genogroupable meningococci n=122 (97.6%) were positive for either vaccine-types menC, menW, menY or non-vaccine types menB, menE, menX and menZ, Compared with a pre-vaccine-implementation baseline, there was 3.8-fold reduction (*p*<0.001) in vaccine-type carriage rates and 9.0-fold increase (*p*<0.0001) in non-vaccine type menE prevalence. These findings imply that menACWY vaccination reduced circulation of vaccine-type meningococci, but lead to serogroup replacement in carriage.

## Introduction

In the Netherlands invasive meningococcal disease (IMD) is a notifiable disease and continuous surveillance of IMD is conducted to evaluate the impact of vaccination and to detect emerging virulent strains. Vaccination against disease caused by *Neisseria meningitidis* was first introduced into the Dutch National Immunization Programme (NIP) in 2002. First used was a conjugated polysaccharide menC vaccine (NeisVac-C, Pfizer) administered at 14-months of age. In response to an outbreak of menW disease, the monovalent menC vaccine for infants was replaced in 2018 with a tetravalent menACWY vaccine (Nimenrix, Pfizer) [1]. To curb the transmission of the hyperinvasive menW clonal complex 11 (cc11) outbreak strain, fifteen-to eighteen-years olds were also vaccinated in a catch-up campaign that started in December 2018 and from 2020 onwards, the NIP was extended with menACWY vaccination at 14 years. The coverage of the vaccine in the Netherlands is currently estimated to be 92.8% and 85.3% for infants and teenagers, respectively [2].

Following the COVID-19 pandemic, a marked decline in IMD was observed in the Netherlands [3]. This could be attributed to the implementation of nationwide non-pharmaceutical interventions (NPIs) and consequent general decline in the circulation of respiratory pathogens, or due to menACWY vaccination [2]. With the reported decline in non-vaccine-type menB IMD [3], the former scenario seems more plausible. To study the effects of menACWY implementation on the circulating meningococci at times of low IMD incidence, monitoring carriage can be considered. While conjugate vaccines are presumed to not only protect against IMD but also confer protection against acquisition of meningococcal carriage, a recent systematic review has reported that insufficient evidence is available for a protective effect of menACWY against vaccine-type carriage [4]. In this carriage study we investigate prevalence of *N. meningitidis* and meningococcal genogroup carriage in Dutch young adults. To assess how menACWY vaccination affects vaccine-type circulation in the population, results from this study were compared to a study conducted in 2018 in the same demographic population and season, which served as a pre-menACWY vaccine introduction baseline of meningococcal carriage and genogroup-specific prevalence rates [5].

## Materials and Methods

### Ethics statement

The study protocol was reviewed by the Centre for Clinical Expertise at the RIVM. Since procedures were considered non-invasive, and participants were anonymized, the study was considered to be outside the scope of the Medical Research Human Subjects Act (http://www.ccmo.nl) and a Medical Ethical Committee issued waiver for additional ethical review. Informed consent was collected from all participants prior to inclusion and the study was conducted in accordance with the World Health Medical Association 1966 Declaration of Helsinki and the EU rules of Good Clinical Practice.

### Study design and sample collection

We have previously reported that the diagnostic performance of saliva to determine meningococcal carriage rates mirrored the diagnostic accuracy of oropharyngeal swabs [5]. Therefore, in this study only saliva was collected and processed (**Fig. 1**) following the protocol established in 2018. In short, saliva was collected from college students of Hogeschool Utrecht (n = 602) in September and October 2022. Students self-collected saliva by spitting approximately 1 ml into a 15 ml tube (Greiner Bio-One B.V, Netherlands). Immediately after collection, 50 μl saliva was used to inoculate Neisseria Selective Medium PLUS agar plates (NS-agar, Oxoid, Badhoevedorp, the Netherlands) and within 20 min plates were placed in a 37°C, 5% CO_2_ incubator. Once all samples were collected, inoculated NS-agar plates were transported at room temperature to the laboratory.

**Figure 1.**
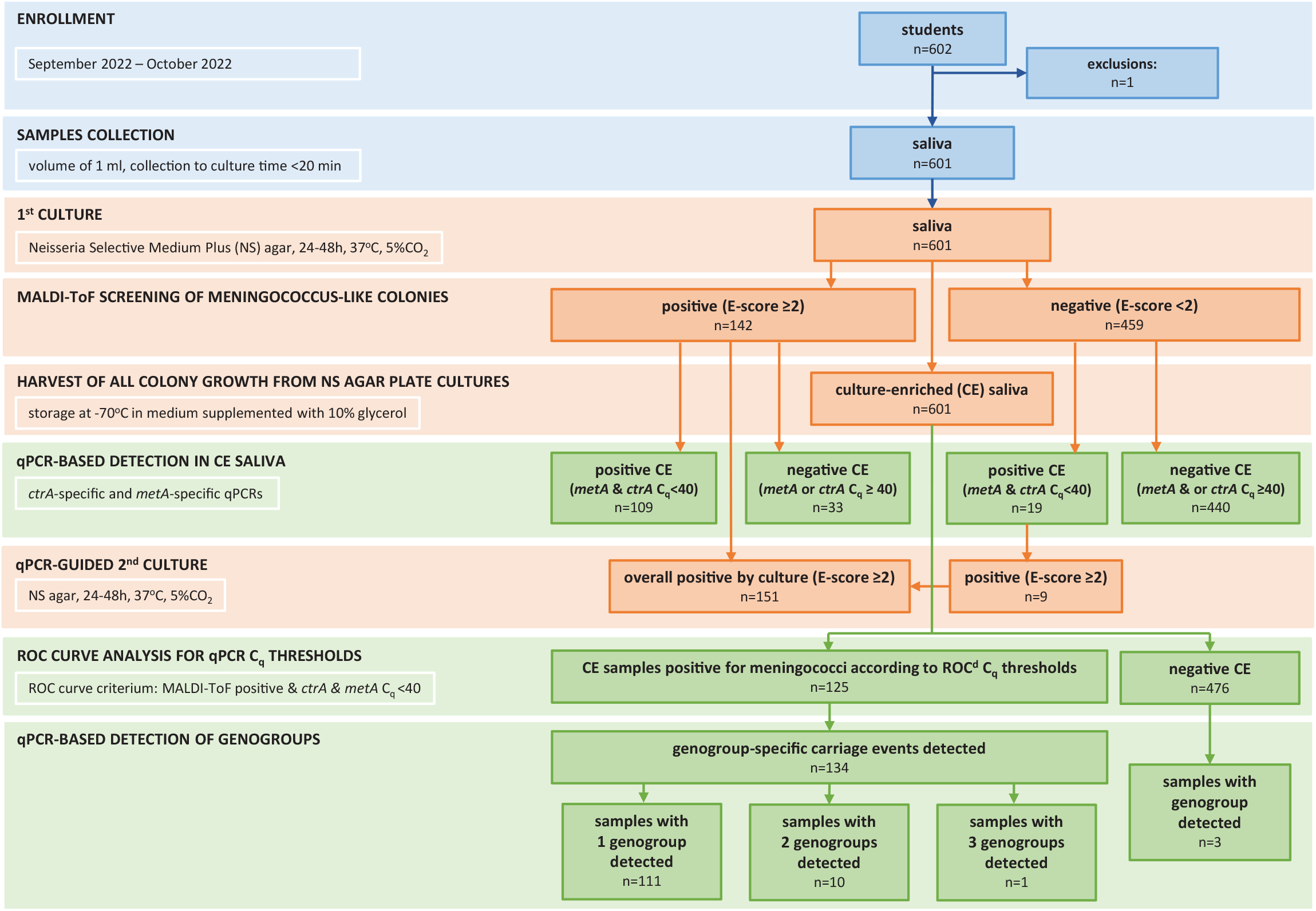
Flowchart depicting the study workflow and results of meningococcal detection using either culture-based or qPCR-based diagnostic methods. Receiver operating characteristic curve analysis was used to determine optimal Cq thresholds using the maximum Youden index and as criterium culture positivity plus quantification of metA and ctrA (<40 Cq) in the culture-enriched saliva sample [7].

### Meningococcal carriage detection using culture

Upon arrival, NS-agar cultures were incubated for two days at 37°C and 5% CO_2_. Thereafter, cultures were screened for presence of meningococcus-like colonies (grey, round and smooth colonies with convex shape). When found, two colonies were re-cultured on Columbia Blood agar (bioTRADING Benelux B.V., Mijdrecht, the Netherlands) and tested for species identification using Matrix-assisted Laser Desorption/Ionization Time-of-Flight mass spectronomy (MALDI-ToF, Bruker Daltonik GmbH, Bremen, Germany). A single isolate with a score ≥ 2.0 for *Neisseria meningitidis* (database BDAL V8.0.0.0 + SR1.0.0.0, Bruker Daltonik) was stored at −70°C in Brain Heart Infusion (Oxoid) supplemented with 0.5% Yeast Extract (YE, Oxoid) and 10% glycerol. NS-agar cultures displaying any microbial growth were harvested into 2 ml of Todd–Hewitt Broth (Oxoid) supplemented with 0.5% YE and 10% glycerol and 1 ml of it stored at −70°C. These harvests were considered to be culture-enriched for meningococci.

### Detection of meningococcal DNA with qPCR

DNA was extracted from 100 μl of harvest of culture-enriched samples using DNeasy Blood and Tissue kit (Qiagen, Hilden, Germany) and eluted in 100 μl volumes as previously described [5]. One microliter of DNA extracts was tested in quantitative-PCRs (qPCRs) using primers and probes (Eurogentec, Seraing, Belgium) targeting sequences within *metA*, a gene encoding for a periplasmic protein, and a capsule transporter gene *ctrA* as described previously [5]. C_q_ thresholds for positivity were determined with receiver operating characteristic (ROC) curve analysis. The specificity of *metA* and *ctrA* qPCR assays were evaluated in Bland-Altman analysis and by calculating intraclass correlation coefficients, as previously described [6].

### Genogroup-specific qPCRs

Following the procedure, primer and probe sequences were used in genogroup-specific qPCR assays targeting genogroups menA, menB, menC, menE, menX, menW, menY or menZ [7], sequences as well as concentrations are listed in **Table S1**. These qPCR assays were used to test culture-enriched samples and were conducted on a LightCycler480, using SensiFast probe No-ROX mastermix (Bioline, London, United Kingdom) and with program described in **Table S2**. Samples were regarded as positive for a genogroup by qPCR when the C_q_ was lower than the cut-off value set for *ctrA*. The specificity of genogroup-specific qPCR assays was determined in Bland-Altman analyses with upper limit of agreement between *metA* and *ctrA* as a priori acceptable limit for genogroup-specific C_q_s [6].

### Statistical analysis

Data was analyzed using Prism (GraphPad Software, v9.3.1) and R (version 4.2.0). ROC curve analysis was performed using “cutpointr” package 25 [5]. Unpaired frequencies were compared with the Fisher’s exact test. To determine the optimal C_q_ thresholds for qPCR detection, maximal Youden indices were determined via bootstrapping (n = 5000) on *metA* and *ctrA* qPCR data with culture as reference. We created Bland-Altman plots with the ‘blandr’ R package [8] and two-way mixed effects intraclass correlation coefficients (ICCs (3,1)) were calculated in a two-way mixed-effects model with consistency relationship (*metA* C_q_ = *ctrA* C_q_ + systematic error) and of single measurement type with the ‘irr’ R package [9]. A *p* value of < 0.05 was considered significant.

## Results and discussion

### Carriage surveillance in young adults

Young adults are reported to be the main reservoir of meningococcus in the population [10]. The age range of sampled individuals was 16-30 (median age of 20) and the proportion of women 59.9% (360 of 601 sampled individuals). The participant characteristics of the current study closely resembled the age and sex distributions of the 2018 study cohort (60.9% female or 182 of 299 sampled individuals; median age 20 years, range 16– 28 years) [5].

Viable meningococci were cultured from 151 saliva samples (25.1% of 601) (**Fig. 1**). Carriage of genogroupable meningococci, was detected with qPCR-based methods in 125 saliva samples (20.8% of 601, **Fig. 2A**). A sample was considered positive for genogroupable meningococci when both *metA* and *ctrA* were detected [5, 7, 11] and we observed high concordance between these two targets (**Fig. 2B**). Next, we conducted genogroup-specific qPCR assays for all vaccine-types plus menB, menE, menX and menZ (**Fig. 3A**). At least one genogroup was detected in 122 of 125 of samples (97.6%) identified as positive for meningococcus by qPCR. Bland-Altman analysis indicated high concordance in quantification of *ctrA* and dominant genogroups (**Fig 3B)**. The most prevailing genogroups were menB and menE, detected in 66 (11.0% of 601) and 36 (6.0%) students, respectively. With qPCR we were able to not only detect dominant, but also minority genogroup carried. Presence of more than one genogroup was detected in samples from 11 students (**Fig. 1**).

**Figure 2.**
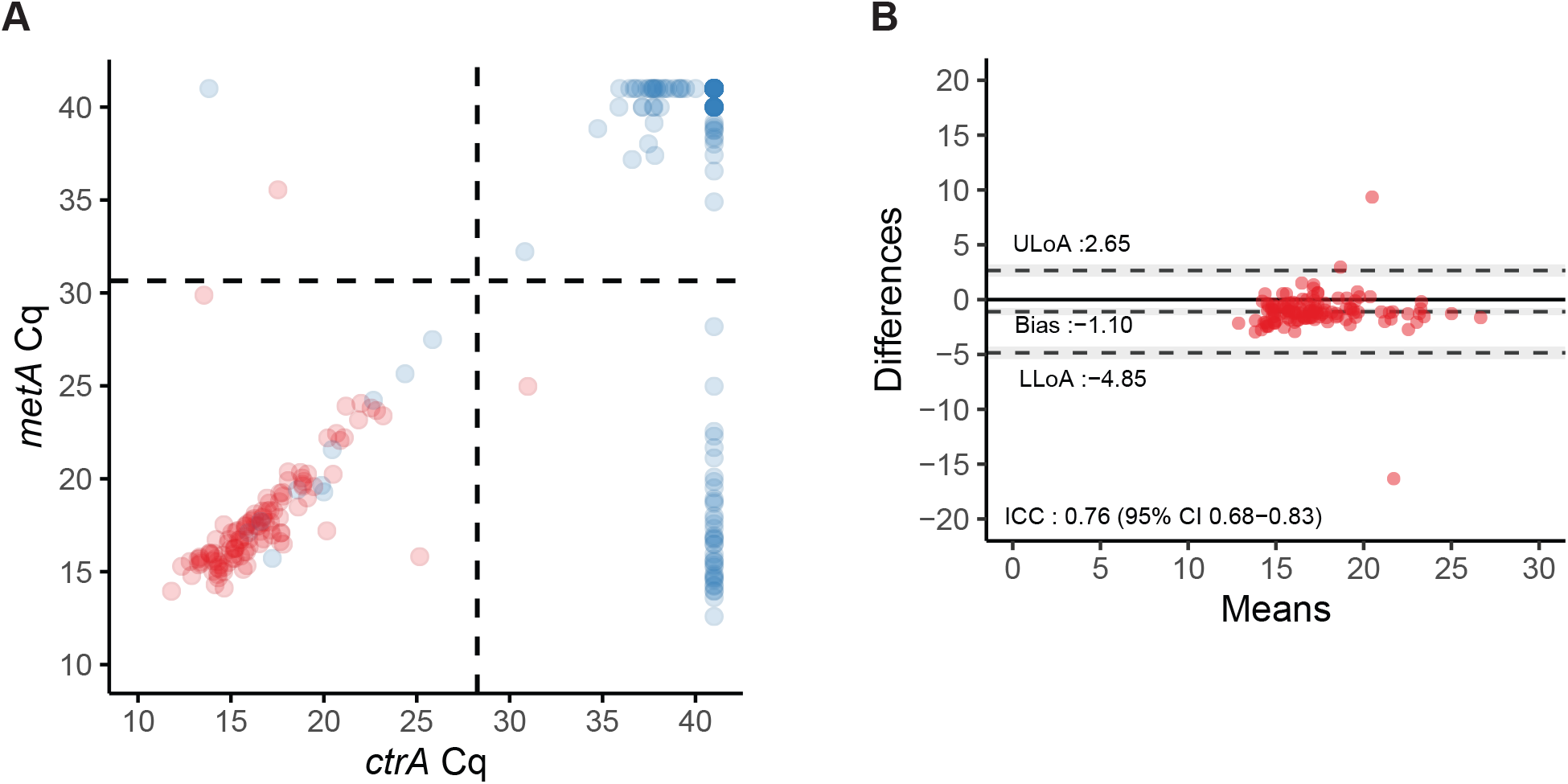
qPCR based detection of *Neisseria meningitidis* in culture-enriched saliva samples. Scatter plot displaying C_q_ values for quantifying ctrA and metA in CE-saliva samples **(A)** and corresponding Bland-Altman plots **(B)** showing extent of agreement and bias among positive samples. Red dots indicate samples positive for culture, blue dots represent culture negative samples. Black dashed lines in scatter plot indicate data-driven thresholds based on receiver operating characteristic curve analysis. In Bland-Altman plots black dashed lines indicate the upper (ULoA) and lower limit of agreement (LLoA) and mean difference (bias). Shaded areas indicate 95% confidence interval. The solid black line indicates the line of equality (no bias). Intraclass correlation coefficient (ICC) <0.50, 0.50-0.75, 0.75-0.90 and >0.90 are indicative of poor, moderate, good, and excellent reliability, respectively.

**Figure 3.**
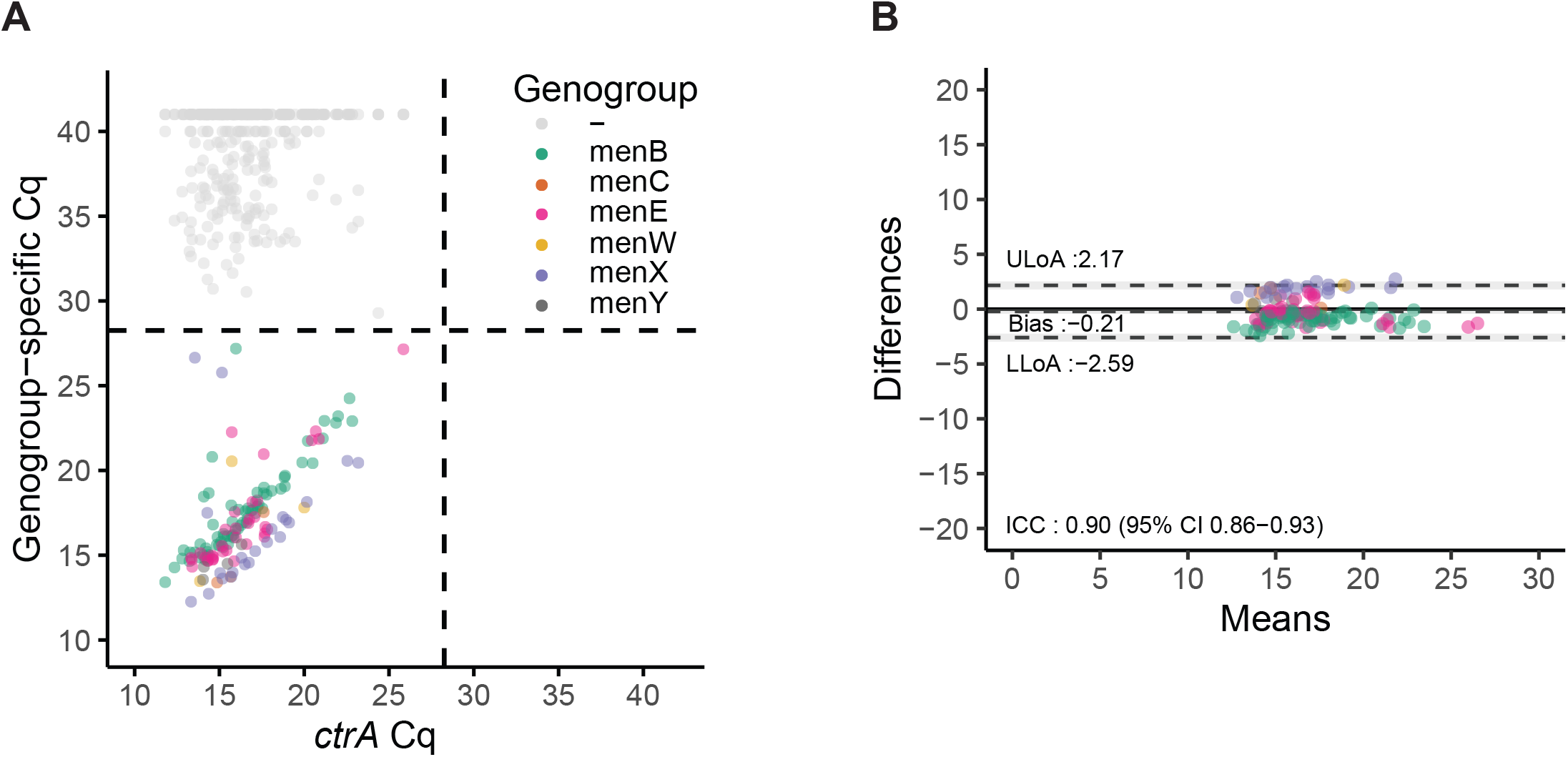
qPCR based genogrouping of *Neisseria meningitidis* positive saliva samples. **(A)** Scatter plot illustrating high agreement between ctrA and genogroup specific qPCRs. **(B)** Bland-Altman plot displays extent of agreement and bias among positive samples. Saliva samples serotype are coloured by genogroup. Black dashed lines in scatter plot indicate data-driven thresholds based on receiver operating characteristic curve analysis analysis. In Bland-Altman plots black dashed lines indicate the upper (ULoA) and lower (LLoA) limit of agreement and mean difference (bias). Shaded areas indicate 95% confidence interval. The solid black line indicates the line of equality (no bias). Intraclass correlation coefficient (ICC) <0.50, 0.50-0.75, 0.75-0.90 and >0.90 are indicative of poor, moderate, good, and excellent reliability, respectively.

### Overall Meningococcal Carriage did not Change after menACWY Vaccine Implementation

Comparing the saliva-based results of the 2018 and 2022 studies allowed us to investigate the impact of menACWY implementation as well as NPIs during the COVID-19 pandemic on meningococcal carriage in young adults. Of note, the 2022 cohort included twice as many students compared to 2018, which was feasible by limiting sampling to saliva. With a prevalence of 17.4% in 2018 and 20.8% in 2022, the carriage rates detected with qPCR in saliva did not differ significantly between studies (Fisher’s exact test, *p=*0.25) (**Fig. 4A**). This is in line with findings by others [12] reporting limited impact of the menACWY vaccine on overall carriage of *N. meningitidis*. Although this observation seems in contrast with a substantial decline in IMD incidence rates during pandemic years [13], it mirrors the dynamics in carriage and disease of another bacterial respiratory pathogen, namely *Streptococcus pneumoniae* [14]. We find insufficient evidence for an impact of NPIs on meningococcal carriage.

**Figure 4.**
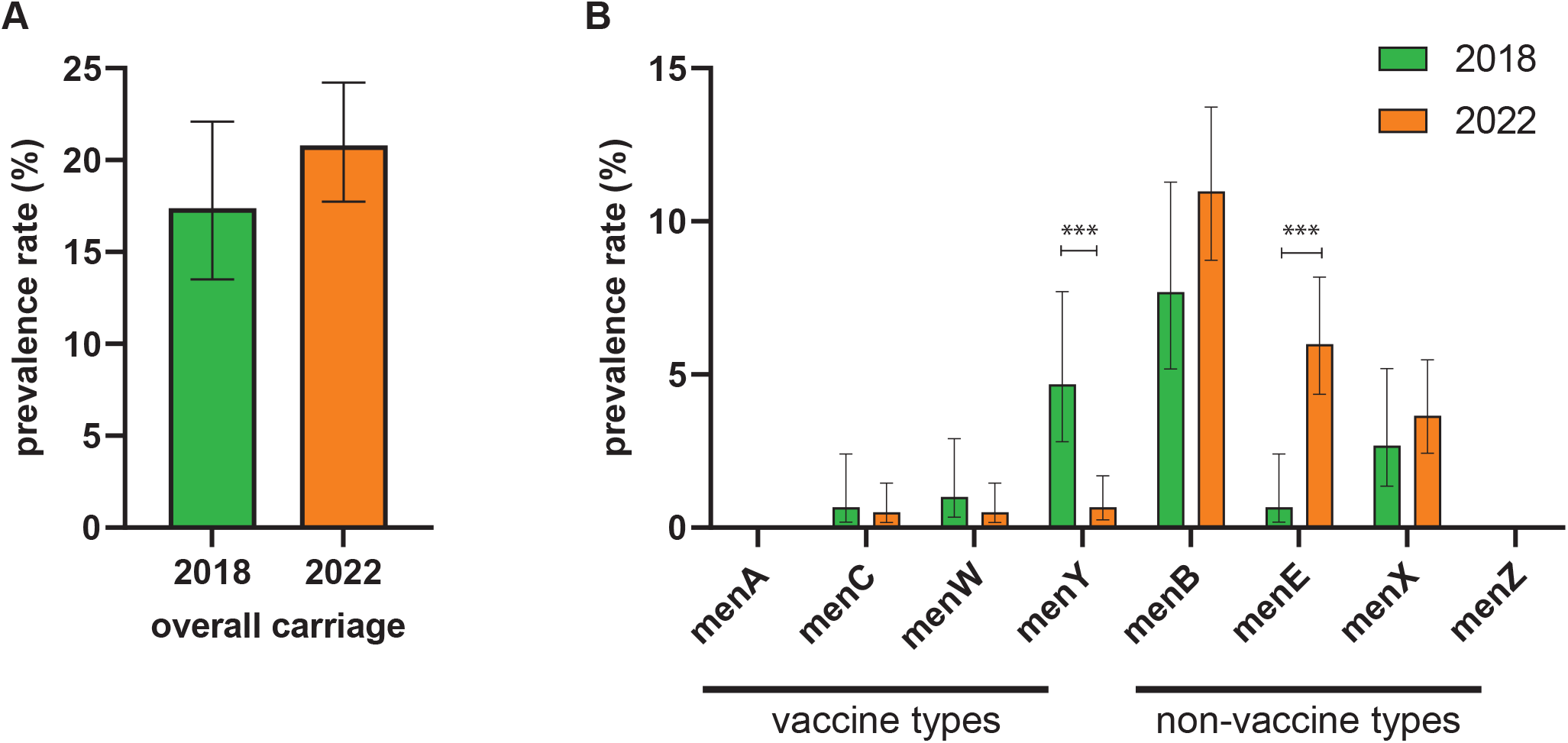
Overall meningococcal **(A)** and genogroup prevalence **(B)** rates among young adults before (2018) and after (2022) the introduction of the MenACWY vaccine in the Netherlands. Data are represented as mean±95%CI. P values were calculated using a Fisher’s Exact test, *** marks *P*<0.001. Of note, samples from the 2018 study have been revisited for menE, menX and menZ carriage since they since they were not tested for these particular genogroups in the original study [7].

### menACWY Vaccination Affected Genogroup Distribution in Carriage

We observed a substantial decline in vaccine-type carriage (**Fig. 4B**), 6.4% to 1.7% (Fisher’s Exact test, *P*<0.001) that amounted to a 3.8-fold reduction between 2018 and 2022 (Fisher’s exact test, *p*<0.001, 0.3 odds ratio, [95%CI 0.1-0.6]). Since baseline rates of menC and menW were low and carriage of menA was altogether absent in the 2018 study, this decline in vaccine-type carriage was mainly driven by a reduction in menY circulation (from 4.7% in 2018 to 0.7% in 2022; Fisher’s Exact test, *P*<0.001). Our findings are in line with a meningococcal carriage study conducted among adolescents in the United Kingdom, where a decline in menACWY vaccine-type prevalence was observed three years after the vaccine was introduced to the national vaccination programme [15]. In the Netherlands, a significant decline in menW but not menY IMD incidence was noted after menACWY implementation in vaccine-noneligible age groups [16]. Collectively, these observations support the notion that menACWY vaccination confers protection against carriage. Importantly, vaccine-type genogroups were not overrepresented among genogroups detected with qPCR as secondary, minority meningococci in co-carriage. Consequently, we did not detect any masked circulation of vaccine-type genogroups and restricting the study to culture-based analyses would not create a bias in proportions of students carrying individual genogroups among all colonized with *N. meningitidis*.

Concurrently, between 2018 and 2022 we observed a 9.0-fold increase in non-vaccine type menE carriage rates (Fisher’s exact test, *p*<0.0001, 9.5 odds ratio [95%CI 2.4-81.6], **Fig. 4B**). Other non-vaccine-type genogroups, including menB, did not exhibit a significant increase in prevalence. IMD surveillance data from the Netherlands indicate near absence of menE disease including zero cases in 2022 (personal communication, N.M. van Sorge and A. Steens) and implies a low invasive potential of menE. This is in line with previous reports describing low menE invasiveness and disease reported primarily in immunocompromised patients [17]. However, considering reports of menE IMD among previously healthy individuals in Queensland Australia caused by a single sequence type strain [18], continued monitoring of circulating menE, is required.

Moreover, genomic-based analyses to assess possible shifts towards more virulent clones among non-vaccine-type serogroups is of importance [19]. Taken together, since overall meningococcal carriage rates remained unaffected, the decline of vaccine-type genogroup carriage can be attributed primarily to menACWY vaccine implementation. Of note, the presence of genogroup-specific sequences does not necessarily translate into colonisation with encapsulated meningococci [20]. Therefore, assessment of capsular locus functionality is warranted.

## Conclusion

In conclusion, the introduction of the menACWY vaccine in 2018 and COVID-19 related restrictions in society seem to have had limited impact on the overall carriage of *N. meningitidis* among young adults, who are considered the main ecological reservoir of meningococci. In the current study diminished vaccine-type circulation was observed after menACWY implementation. This finding implies that menACWY vaccination prevents carriage of vaccine serogroups. Strikingly, we observed a rise in menE carriage prevalence, suggesting serogroup replacement in meningococcal carriage.

## Data Availability

All data produced in the present study are available upon reasonable request to the authors

## Acknowledgements

We gratefully acknowledge the students of Hogeschool Utrecht for their participation in the study. We also wish to thank Janieke van Veldhuizen and Tessa Nieuwenhuijsen for the technical assistance in the lab, and Dr. Anneke Steens for valuable discussion of the results presented in the paper.

