## Supplemental Material for "Surveillance of *Neisseria meningitidis* Carriage Four Years After menACWY Vaccine Implementation in the Netherlands Reveals Decline in Vaccine-type and Rise in Genogroup E Circulation"

| **Oligonucleotide** | **Sequence** | **Concentration (nM)** | **Reference** |
| --- | --- | --- | --- |
| *metA* forward primer | 5'-GCGAATTTGCTAATCCTATTTATGTGC-3' | 750 | Diene *et al* 2016 |
| *metA* reverse primer | 5'-AAATTTTGCGCCATTACAGGTG-3' | 750 |  |
| *metA* probe | 5'-6FAM-AACCAGCGCAACGAAAATTGCAA-3'-TAMRA | 200 |  |
| *ctrA* forward primer | 5'-TGGGCGGTTTGCAAGATC-3' | 500 | Rojas *et al* 2015 |
| *ctrA* reverse primer | 5'-TGACGTTCTGCCGGCAAT-3' | 500 |  |
| *ctrA* probe | 5'-6-FAM-CACACCACGCGCATCA -3'-TAMRA | 200 |  |
| genogroup A forward primer | 5’-GCCACAAAGTGCCCTTCCT-3’ | 800 |  |
| genogroup A forward primer | 5’-TGGTATATGGTGCAAGCTGGTT-3’ | 800 |  |
| genogroup A probe | 5’-6-FAM-TTTAGCTCACATGCTATTG-3’-TAMRA | 300 |  |
| genogroup B forward primer | 5’-CCTCGGCTGGTAGTTATTAATGAAC-3’ | 300 |  |
| genogroup B reverse primer | 5’-GCCAGGCCTATAATTCCTTTAGGA-3’ | 300 |  |
| genogroup B probe | 5’-6-FAM-CCTTTTCTAATTGAGCCCCTAA-3’-TAMRA | 100 |  |
| genogroup C forward primer | 5’-GCACATTCAGGCGGGATTA-3’ | 200 |  |
| genogroup C reverse primer | 5’-TTGAGATATGCGGTATTTGTCTTGA-3’ | 100 |  |
| genogroup C probe | 5’-6-FAM-ACAAGCCAATCTATTGCT-3’-TAMRA | 400 |  |
| genogroup E forward primer | 5’-AGATTTACGTCGTGCCTTCGA-3’ | 400 |  |
| genogroup E reverse primer | 5’- CATGCATATAGGCTTCAGCTTCTG-3’ | 400 |  |
| genogroup E probe | 5’- FAM-TCCTAAATATGTTTCTGCTGATGCCCGC-3’-TAMRA | 200 |  |
| genogroup X forward primer | 5’-TTGCGCCGCCATAAAGA-3’ | 400 |  |
| genogroup X reverse primer | 5’-GAGGCAGATTTGCGTTTTGG-3’ | 400 |  |
| genogroup X probe | 5’-FAM-ACTGACACCACCTTC-3’-MGB | 100 |  |
| genogroup Y forward primer | 5’-GTACGATATCCCTATCCTTGCCTATAA-3’ | 200 |  |
| genogroup Y reverse primer | 5’-CCATTCCAGAAATATCACCAGTTTTA-3’ | 100 |  |
| genogroup Y probe | 5’-6-FAM-TGGAGCGAATGATTTTAGCAA-3’-TAMRA | 100 |  |
| genogroup Z forward primer | 5’-GCACTATGGTCATATTCGCCTTT-3’ | 400 |  |
| genogroup Z reverse primer | 5’-TCATCAGTTGAGATGGCAACAGT-3’ | 400 |  |
| genogroup Z probe | 5’-FAM-AACGTGCAAAAGCTCTAGGCGACCATC-3’-TAMRA | 200 |  |
| genogroup W forward primer | 5’-CAGAAAGTGAGGGATTTCCATA-3’ | 200 | Knol *et al* 2017 |
| genogroup W reverse primer | 5’-CACAACCATTTTCATTATAGTTACTGT-3’ | 100 |  |
| genogroup W probe | 5’-6-FAM-TGGAAGGCATGGTGTATGATATTC-3’-TAMRA | 100 |  |

| **qPCR assay** | **Step** | **Cycles** | **Temperature (°C)** | **Duration** |
| --- | --- | --- | --- | --- |
|  | Pre-incubation | 1 | 95 | 10 min |
| *metA* and *ctrA* qPCR | Denaturation |  | 95 | 10 sec |
|  | Annealing | 45 | 60 | 45 sec |
|  | Elongation |  | 72 | 1 sec |
|  | Pre-incubation | 1 | 95 | 5 min |
| serogroup-specific qPCR | Denaturation | 45 | 95 | 10 sec |
| Annealing | |  | 60 | 50 sec |
| Elongation | |  | 72 | 1 sec |
